# Quantifying the bystander effect of antimicrobial use on the diversity and resistome of the gut microbiome in Malawian adults

**DOI:** 10.1101/2025.04.03.25325172

**Authors:** Edward Cunningham-Oakes, Vivien Price, Madalitso Mphasa, Jane Mallewa, Alistair Darby, Nicholas A Feasey, Joseph M Lewis

## Abstract

Antibiotic treatment for sepsis has an unintended yet crucial consequence: it exerts a bystander effect on the microbiome, changing its bacterial composition and resistome. Antimicrobial stewardship aims, in part, to minimise this effect to prevent development of subsequent drug-resistant infection, but data evaluating and quantifying these changes are largely lacking, especially in low-income settings which are disproportionately affected by antimicrobial resistance. Such data are critical to creating evidence-based stewardship protocols.

We address this data gap in Blantyre, Malawi. We used longitudinal sampling of human stool and metagenomic deep sequencing to describe microbiome composition and resistome pre-, during- and post-antimicrobial exposure. We developed Bayesian regression models to link these changes to individual antimicrobial agents. We find that ceftriaxone, in particular, exerts strong off-target effects, both increasing abundance of Enterobacterales, and the prevalence of macrolide and aminoglycoside resistance genes. Simulation from the fitted models allows exploration of different stewardship strategies, and can inform practice in Malawi and elsewhere.

## Introduction

Antimicrobial resistance (AMR) is a global public health threat, and a key strategy to prevent it is antimicrobial stewardship. Stewardship aims to avoid unnecessary antimicrobials, to reduce duration of antimicrobial use to the minimum necessary, and to select an antimicrobial agent with as narrow a spectrum of antibacterial activity as possible^1^. These principles aim in part to minimise antimicrobial pressure for promotion of AMR in bacteria other than the intended pathogenic target - the so-called bystander effect^2^. Even though minimising the bystander effect is a key factor in selection of an antimicrobial agent for a given infection, data quantifying the magnitude and duration of this effect for a given antimicrobial at the individual level are lacking.

Addressing this is key to designing and implementing antimicrobial strategies that can minimise development of AMR. In sepsis, for example, it is recognised that early antimicrobials confer a survival advantage and global efforts to improve sepsis care prioritise early antimicrobials in suspected sepsis^3^. But this strategy may result in widespread administration of broad-spectrum antimicrobials, including to people who are ultimately found not to have sepsis or an infectious cause of their illness. This is particularly the case in low-resource settings where diagnostics to inform targeted antimicrobial treatment are frequently unavailable. To truly assess the impact of such a strategy requires not only descriptions of clinical outcomes but tools to quantify unintended effect of antimicrobials in promoting AMR, both in pathogens and in bystander bacteria.

Shotgun metagenomic sequencing provides a method to achieve this, enabling a quantitative assessment of bacterial taxa in a sample (microbiota) and antimicrobial resistance genes (resistome), and linking changes to antimicrobial exposures. Most work has focussed on the development of the gut microbiome in the first days to years of life, where antimicrobial exposure delays development of a mature microbiome^4^, reduces diversity^5^ and promotes colonisation with resistant organisms^6,7^, with evidence that narrower-spectrum treatment may reduce this effect^8^. In adults, data are largely restricted to healthy volunteers^9,10^ or specific cohorts^11^ (e.g. inflammatory bowel disease). However, data from cohorts of adults with severe febrile illness, particularly from LMIC - the cohorts who may be expected to have a significant broad-spectrum antimicrobial exposure - are lacking, as are analytic approaches to attribute bystander effects to individual antimicrobial agents.

We address this data gap here, leveraging our prior study which described aetiology and clinical outcomes of a cohort of adults admitted to hospital in Blantyre, Malawi with sepsis^12^. In that study we described development of gut mucosal colonisation with Extended-spectrum beta-lactamase producing Enterobacterales (ESBL-E), as defined by longitudinal sampling and selective culture^13^. We linked antimicrobial exposure to ESBL-E carriage with a Bayesian modelling approach, demonstrating that antimicrobial exposure acted with a prolonged effect to promote ESBL-E carriage. Here, we build on this: we aimed to describe the dynamics of gut microbiota and resistome of the cohort under antimicrobial pressure, expanding our models to quantify the bystander effect of individual antimicrobial agents.

## Results

### Quantifying the bystander effect on microbiome: ceftriaxone and ciprofloxacin exposure is associated with increased Enterobacterales abundance

The original study recruited 425 adults between February 2017 and October 2018 into three study arms: 225 participants with sepsis admitted to Queen Elizabeth Central Hospital, Blantyre Malawi, and exposed to antimicrobials, 100 age- and sex-matched antimicrobial-unexposed hospital inpatients and 100 community members. Stool or rectal swab samples were collected at recruitment then at four subsequent visits: 7, 28, 90 and 180 days later, except for community members who did not have day 28 or 90 samples. For the current analysis we carried out shotgun metagenomic sequencing on a subset of samples: 426 samples from 162 participants passed QC and were incorporated. Table 1 shows demographics of included participants. Most of the cohort (109/162, 67%) received at least one antimicrobial, most commonly ceftriaxone (94/109 of antimicrobial-exposed, 86%), followed by co-trimoxazole (56/109, 51%), ciprofloxacin (30/109, 28%) and amoxicillin (25/162, 15%, Supplementary Table 1). Cotrimoxazole had a prolonged course length compared to other antimicrobials (Supplementary Table 1) because use was commonly (52/56 cotrimoxazole exposure, 93%) as preventative therapy in the context of HIV; this also had a reduced dose compared to treatment (480mg once daily versus 960mg twice daily).

**Table 1:**
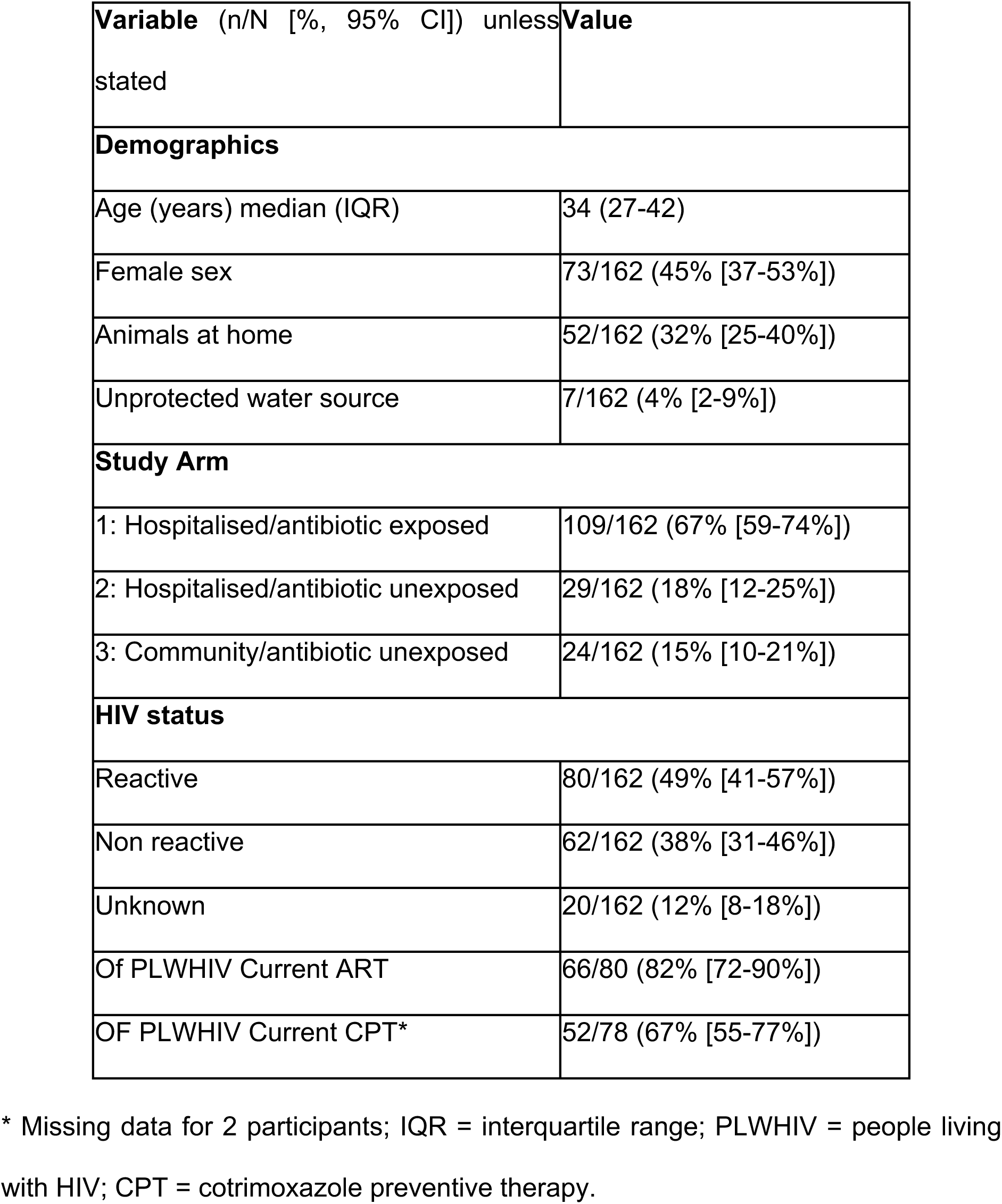
Characteristics of included participants.

| **Variable** (n/N [% , 95% CI]) unless stated | **Value** |
| --- | --- |
| **Demographics** |  |
| Age (years) median (IQR) | 34 (27-42) |
| Female sex | 73/162 (45% [37-53%]) |
| Animals at home | 52/162 (32% [25-40%]) |
| Unprotected water source | 7/162 (4% [2-9%]) |
| **Study Arm** |  |
| 1: Hospitalised/antibiotic exposed | 109/162 (67% [59-74%]) |
| 2: Hospitalised/antibiotic unexposed | 29/162 (18% [12-25%]) |
| 3: Community/antibiotic unexposed | 24/162 (15% [10-21%]) |
| **HIV status** |  |
| Reactive | 80/162 (49% [41-57%]) |
| Non reactive | 62/162 (38% [31-46%]) |
| Unknown | 20/162 (12% [8-18%]) |
| Of PLWHIV Current ART | 66/80 (82% [72-90%]) |
| OF PLWHIV Current CPT* | 52/78 (67% [55-77%]) |

\* Missing data for 2 participants; IQR = interquartile range; PLWHIV = people living with HIV; CPT = cotrimoxazole preventive therapy.

We first examined microbiome composition stratified by study arm and visit. There were shifts in alpha diversity (Shannon diversity) most marked at visit 1 (day 7) in the hospitalised/antimicrobial exposed group (Figure 1A), corresponding to the time of maximal antimicrobial exposure. We explored this effect with linear modelling of Shannon diversity. A within-participant correlation structure accounted for repeated sampling, and we included the top four antimicrobials - ceftriaxone, ciprofloxacin, cotrimoxazole and amoxicillin and hospitalisation as covariates, with an exponential decay of effect following cessation. This identified that ceftriaxone exposure (Fig 1B, 1D) was most strongly associated with changes in diversity; cotrimoxazole had the least effect. Hospitalisation itself was also associated with reduction in diversity (Fig 1B,D). Shannon diversity returned to baseline by ∼ 50 days in simulations following perturbation (Fig 1D).

**Figure 1:**
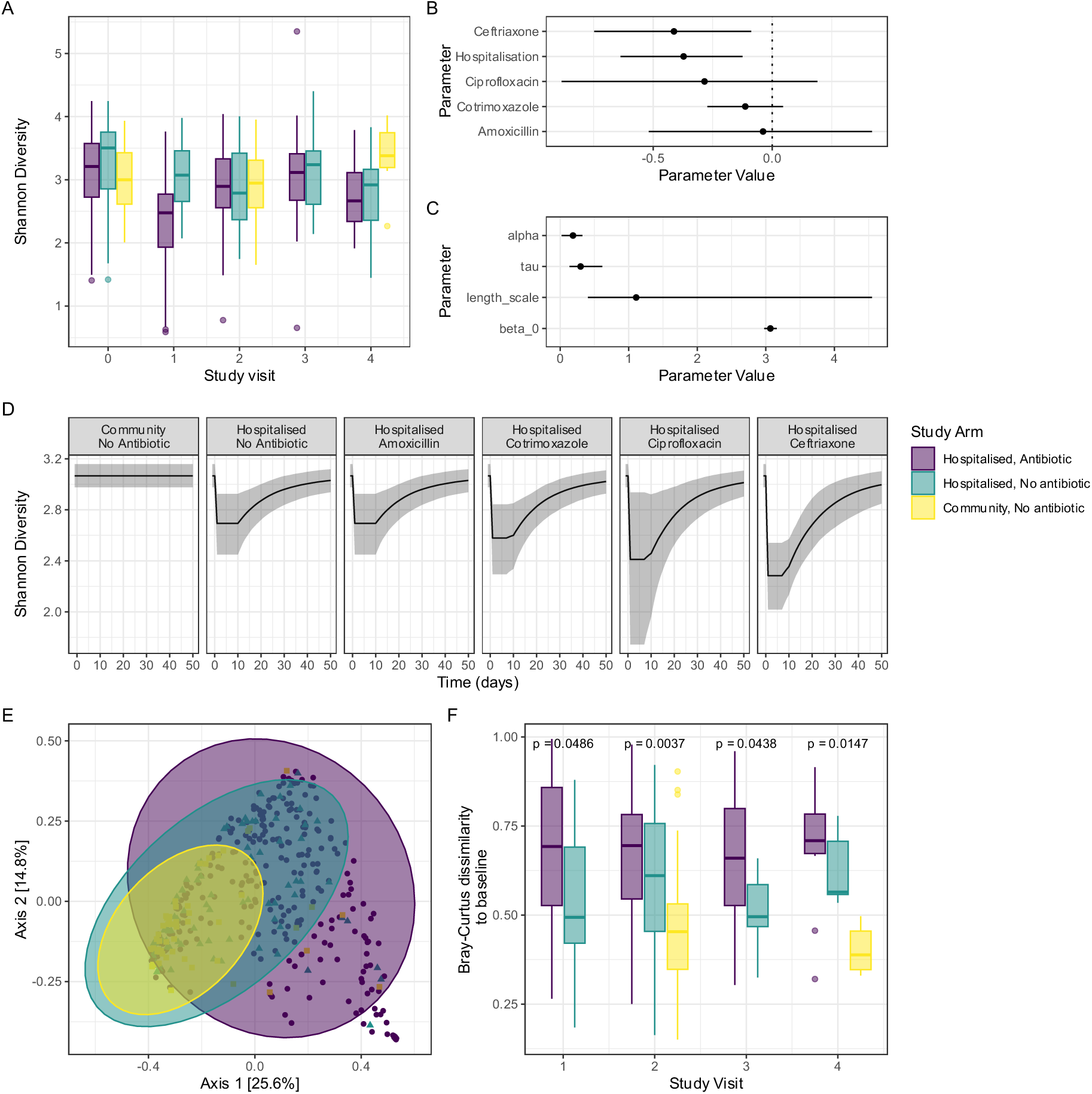
Changes in alpha and beta diversity over time; (A) Shannon Diversity stratified by study arm and visit; (B-C) parameters from modelling Shannon Diversity as a function of antimicrobial exposure. Alpha = magnitude of within-participant correlation, length scale = decay parameter of within participant correlation (on standardised time scale, 1 unit = 55 days), beta_0 = model population intercept, tau = decay constant of effect of antimicrobial exposure (on standardised time scale, 1 unit = 55 days). Antimicrobial/hospitalisation parameters can be interpreted as the change in mean Shannon Diversity given exposure; (D) simulated mean Shannon Diversity for hospitalisation (10 days) and antimicrobial exposure (7days) with different antimicrobials; (E) Principal coordinate plot of all-against-all Bray Curtis dissimilarity (beta diversity) with 95% confidence intervals assuming student T distribution stratified by study arm showing between-arm differences in beta diversity; (F) Within-participant Bray-Curtis dissimilarity to baseline sample, stratified by arm, showing that participants admitted to hospital and exposed to antimicrobials have persistent changes in beta diversity over six months, compared to community and hospital controls. P values are from a Kruskall-Wallace test between all groups at a given visit.

We assessed beta diversity with all-against-all Bray-Curtis dissimilarity and principal components analysis; there was overlap of all three arms of the study in PCA space, but some hospitalised/antimicrobial exposed participants fell outside the distribution of the antimicrobial unexposed participants, suggesting different microbiome composition (Figure 1E). Hospitalised/antimicrobial exposed participants had persistent differences in Bray-Curtis dissimilarity between baseline and subsequent samples in within-participant (Figure 1E), compared to controls. Hence, though Shannon diversity returned to baseline, there is some evidence for changes in microbiome composition due to antimicrobial exposure, which persist out to 6 months following exposure.

At all time points and across all arms the most abundant Phylum was Bacteroidetes (Figure 2A); Proteobacteria (largely Enterobacterales) were more abundant at visit 1 (day 7) in the hospitalised/antimicrobial exposed group (Figure 2A-C), the time point which corresponds to maximal antimicrobial exposure. To quantify this, we fit negative-binomial Bayesian mixed effects models to the absolute number of reads assigned to a given taxon with a per-participant random effect with a multivariate-normal correlation structure to account for repeated measurements, a per-sample read depth offset to account for varying sampling depth and covariates as above. A separate model was fit for each of the top three phyla, the top-10 orders of phylum Proteobacteria, and the top-10 genera of order Enterobacterales.

**Figure 2:**
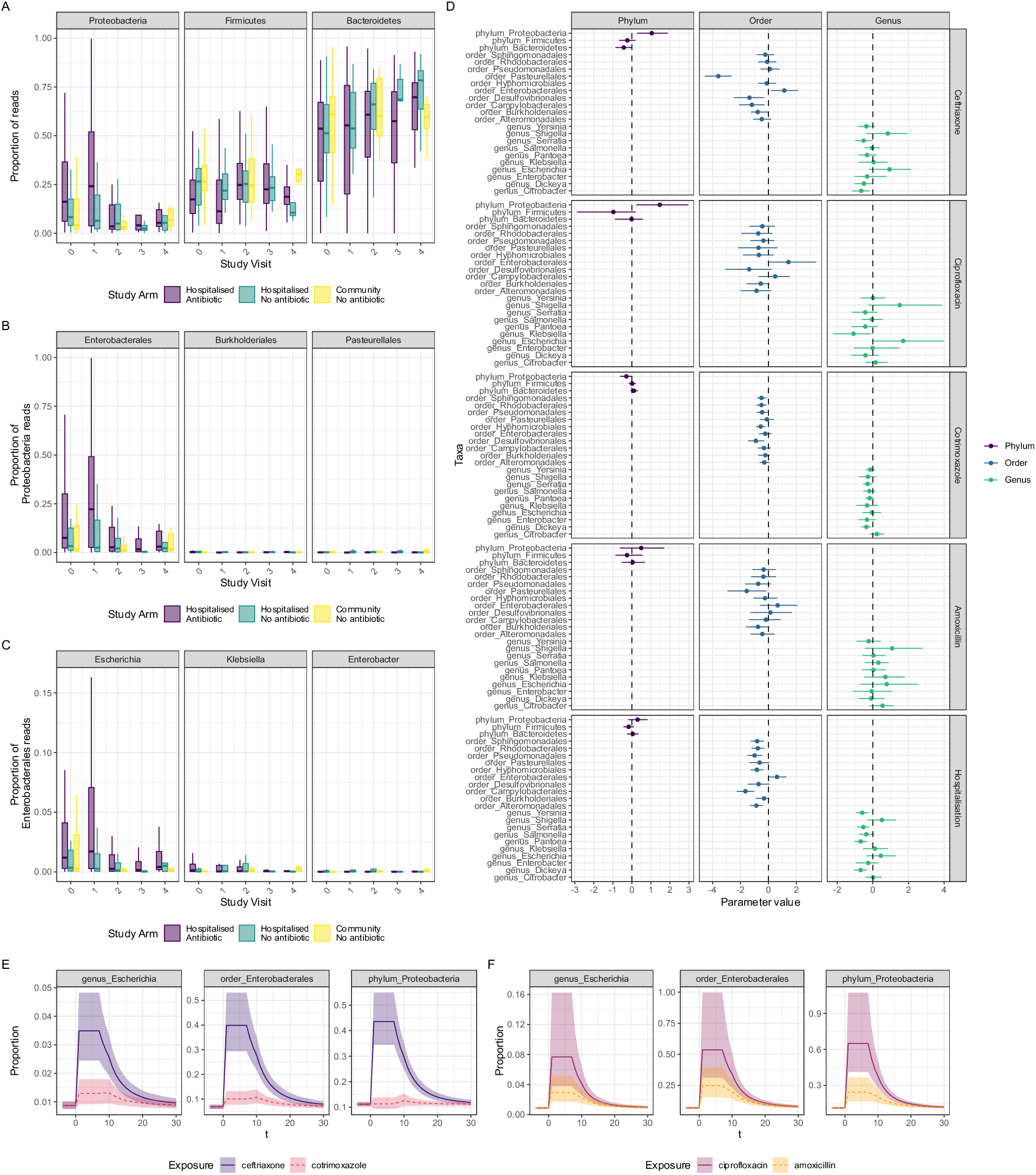
Changes in microbiome composition under antimicrobial pressure: Relative abundance of top 3 Phyla (A), top 3 Proteobacteria Orders (B) and top 3 Enterobacterales Genera (C), stratified by study arm and visit, showing higher abundance of Proteobacteria, Enterobacterales and Escherichia at visit 1 in the antibiotic exposed, corresponding to maximal antimicrobial exposure. (D) parameter values for effect of antimicrobial exposure or hospitalisation on microbiome composition, on log scale with 95% credible intervals; parameter values > 1 correspond to an increase < 1 to a decrease. (E-F) simulated antimicrobial exposures (7 days) showing proportion of Proteobacteria reads (right panel), proportion of Proteobacteria reads that are Enterobacterales (middle panel), proportion of Enterobacterales reads that are Escherichia (left panel) with 50% prediction intervals.

The results of these models are shown in Figure 2 and Supplementary Figure 1. Ceftriaxone and ciprofloxacin are convincingly associated (95% CrI effect > 1 on log scale) with increase in Proteobacteria abundance, driven by increase in Order Enterobacterales and, within that order, the genera Escherichia and Shigella (though confidence intervals on genus-level parameter estimates are wide and 95% CrI cross 1). The exposures acted with a prolonged effect: across all models, the posterior median half-life of the exponential decay of exposure effect was 9 (range 11-158) days (Supplementary Figure 1). Simulation of the fitted models (Figure 2E-F) allows comparison of the effect of different antimicrobial strategies on microbiome composition; generally, antimicrobial-associated effects return to baseline by 30 days following exposure.

### Ceftriaxone exerts a bystander effect on the resistome, apparently driving prevalence of aminoglycoside and macrolide resistance

We used metagenome assembly followed by AMRFinderPlus plus to define each individual’s resistome over time. Across the 426 samples, we identified 369 unique AMR genes of 25 classes and 60 subclasses (Figure 2A, Supplementary Figure 2); trimethoprim, tetracycline, beta-lactam, sulphonamide, aminoglycoside and lincosamide/macrolide/streptogramin resistance genes were present in almost all samples. The plasmid-mediated colistin resistance gene *mcr* was detected twice (*mcr10* and *mcr10.1*): in two different participants, neither of whom had hospital admission within the previous 3 months, despite colistin being prohibited in Malawi, as seen in a recent large *E. coli* genome collection from Blantyre^14^. Many of the cephalosporin and carbapenem resistance genes were Bacteroidetes-specific and less clinically relevant in terms of causing drug-resistant infection in our setting (Supplementary Figure 3); excluding these (which we defined as defined as *cfiA, cblA, crxA, cepA* or *cfxA* beta-lactamases), cephalosporin and carbapenem resistance genes were identified in 74% (102/138) and 7% (10/138) of samples from hospitalised participants respectively (Supplementary Table 2), most commonly *bla*_CTX-M-15_ and *bla*_OXA-181_ (Supplementary Figure 3).

Some gene subclasses were more prevalent in hospitalised/antimicrobial exposed group at visit 1 (day 7) than other study arms or visits, corresponding to time of maximal antimicrobial exposure (Supplementary Figure 4): those conferring aminoglycoside, macrolide and cephalosporin resistance, consistent with these subclasses being associated with antimicrobial exposure. To quantify this effect and relate it to individual antimicrobial agents we fit models identical to the taxonomy models but including presence/absence of AMRFinderPlus gene subclass in a mixed-effect logistic regression model (Figure 2, Supplementary Figures 5 and 6). These confirmed that ceftriaxone exposure was convincingly associated (95% CrI of odds ratio >1) with subsequent presence of genes conferring resistance to aminoglycosides (primarily *aac(6’)-i, aac(3”)-ii* and *aph(3’)-ii* alleles), macrolides (*mphA*, *msrC, ermB*) and rifamycins (*arr*). The effect of ciprofloxacin was similar: it was associated with presence of aminoglycoside (*aac(6’)-i*) and macrolide (*mphA, ermC*) genes. Amoxicillin and cotrimoxazole were convincingly associated with fewer resistance subclasses than ceftriaxone (amoxicillin with *mphA* macrolide resistance and cotrimoxazole with *msrD* and *mefA* macrolide resistance and *qnr* plasmid-mediated quinolone resistance).

The 95% CrI of effect estimates for association of all antibiotics with cephalosporin resistance genes crossed the null (Figure 2C)), but cephalosporin resistance genes were very commonly detected across all samples, as Bacteroides-specific genes were included in this analysis (Figure 2B) which could attenuate an effect in other Genera. Hence we stratified all beta lactamases as Bacteroides-associated (defined as *cfiA, cblA, crxA, cepA* or *cfxA* beta-lactamases) or not (all other beta-lactamases), and refit the models (Figure 3F). This showed that, consistent with our previous culture-based analysis, there was an association of ceftriaxone exposure with presence of non-Bacteroides cephalosporin resistance genes (largely *bla*_CTX-M-15_); hospitalisation and cotrimoxazole exposure also showed an association with cephalosporin resistance, as did amoxicillin and ciprofloxacin, though in the case of the latter two antimicrobials the 95% CrI crossed the null. Cotrimoxazole was also associated with presence of narrow spectrum beta lactamases (most commonly *bla*_TEM_ and *bla*_OXA_)

**Figure 3:**
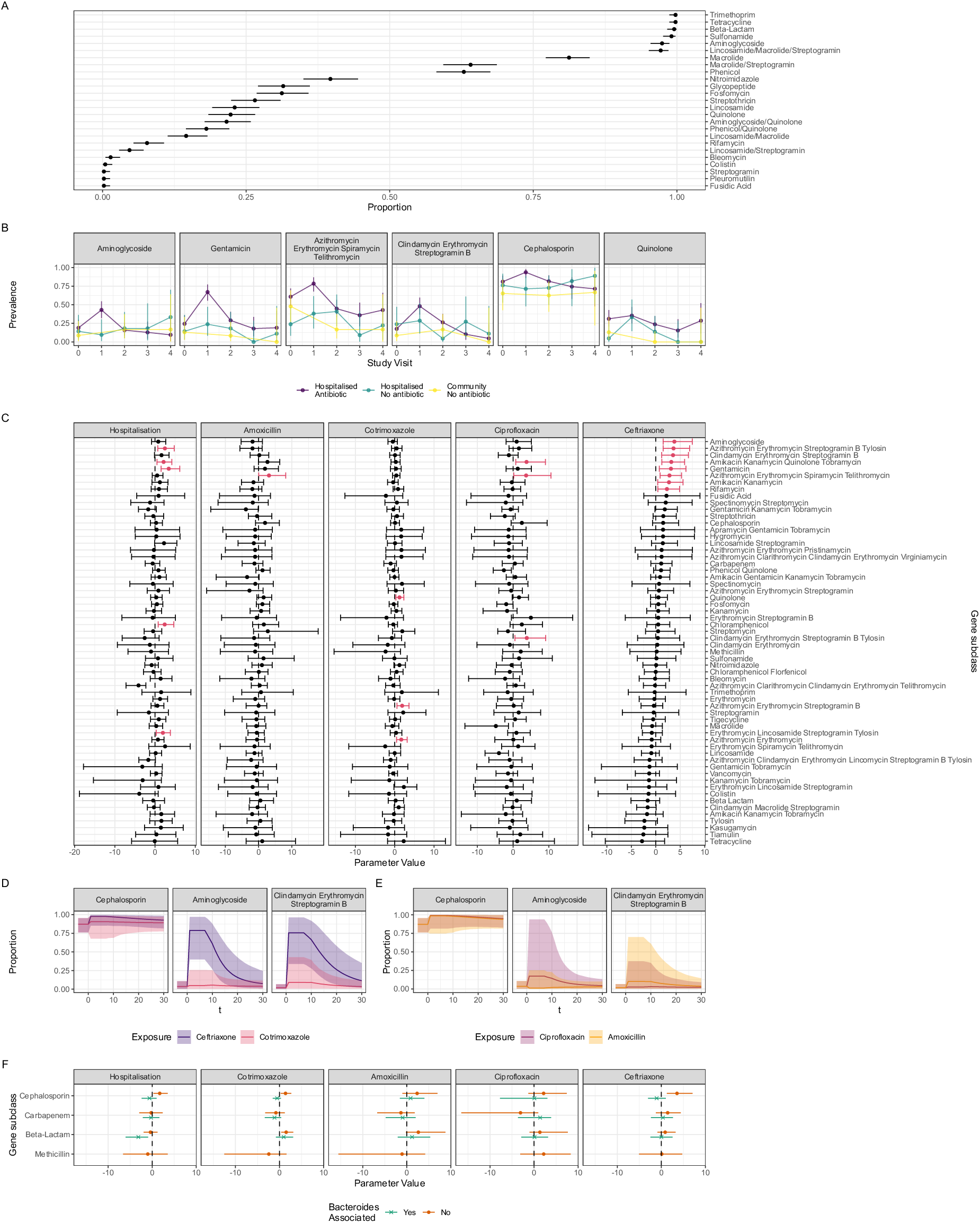
Changes in resistome following antimicrobial exposure. (A) Prevalence of AMRFinderPlus-defined resistance gene subclass. (B) Selected AMRFinderPlus-defined gene subclass prevalence stratified by visit and arm. Cephalosporin and quinolone resistance gene prevalence shows little relationship with visit/arm but macrolide and aminoglycoside resistance subclasses have a higher prevalence at visit 1 in the hospitalised/antimicrobial exposed group, consistent with an association with antimicrobial exposure. (C) Parameter values from modelling antimicrobial resistance gene subclass presence as a function of antimicrobial exposure. Parameter values (and 95% CrI) can be interpreted as logged odds ratio. Parameter values with a convincing association between exposure and outcome (defined at lower bound of 95% CrI > 0) are coloured red. (D-E) simulated AMR gene subclass prevalence for selected subclasses, following a 10-day hospital admission and 7-day exposure to a given antimicrobial agent. Lines show median posterior prediction, shaded area 95% quantile. Ceftriaxone is associated with an increase in aminoglycoside and clindamycin-erythromycin-streptogramin B subclass genes not seen with the other agents. (F) Associations of exposures to presence of AMR gene subclass restricted to beta-lactamases with models fit separately to Bacteroides-associated genes (defined as *cfiA, cblA, crxA, cepA* or *cfxA* beta-lactamases) or all others (defined as all other genes).

Overall, antimicrobials acted with a prolonged effect (across all models posterior median (range) of half-life 11 (3-159) days); simulations from the posterior quantify the profound effect of ceftriaxone in particular in driving aminoglycoside and macrolide resistance genes (Figure 2D-E), and again show that antibiotic-induced perturbations in resistome return to baseline by around 30 days.

### Resistance is associated with clinically relevant Enterobacterales, including *E. **coli***

Presence of genes of the macrolide and aminoglycoside resistance subclasses (as well as chloramphenicol and the fluoroquinolone subclasses) correlated with Proteobacteria abundance (Supplementary Figure 7), particularly Enterobacterales, and Escherichia within this order. To explicitly link AMR gene presence to clinically relevant Enterobacterales, we focussed on *E. coli*. We binned all assembled contigs, identified bins comprising *E. coli* (defined as average nucleotide identity to an E. coli reference > 95%) and identified AMR genes that could be attributed to *E. coli* using AMRFinderPlus using the *E. coli* specific models which allowed identification of point mutations conferring resistance (e.g. in quinolone-resistance determining region, QRDR) as well as AMR gene presence. We compared the diversity of the genomes identified using this approach to our previous analysis of *E. coli* genomes we identified in the same samples using ESBL-selective media by clustering the genomes using popPUNK v2.7.2. As expected, the genomes from the metagenomic approach were more diverse (33 unique popPUNK clusters in culture-derived genomes versus 95 in the metagenome-derived genomes, Figure 4A). We identified 93 unique AMR associated genes/mutations in 260 *E. coli* bins: predicted fluoroquinolone resistance was common (128/260 [49%] samples, commonly *gyrA* and *parC* mutations), as was trimethoprim (75/260 [29%], all *dfr* alleles) and cephalosporin resistance (68/260 [26%], most commonly *bla*_CTX-M-15_ Supplementary Figure 8).

**Figure 4:**
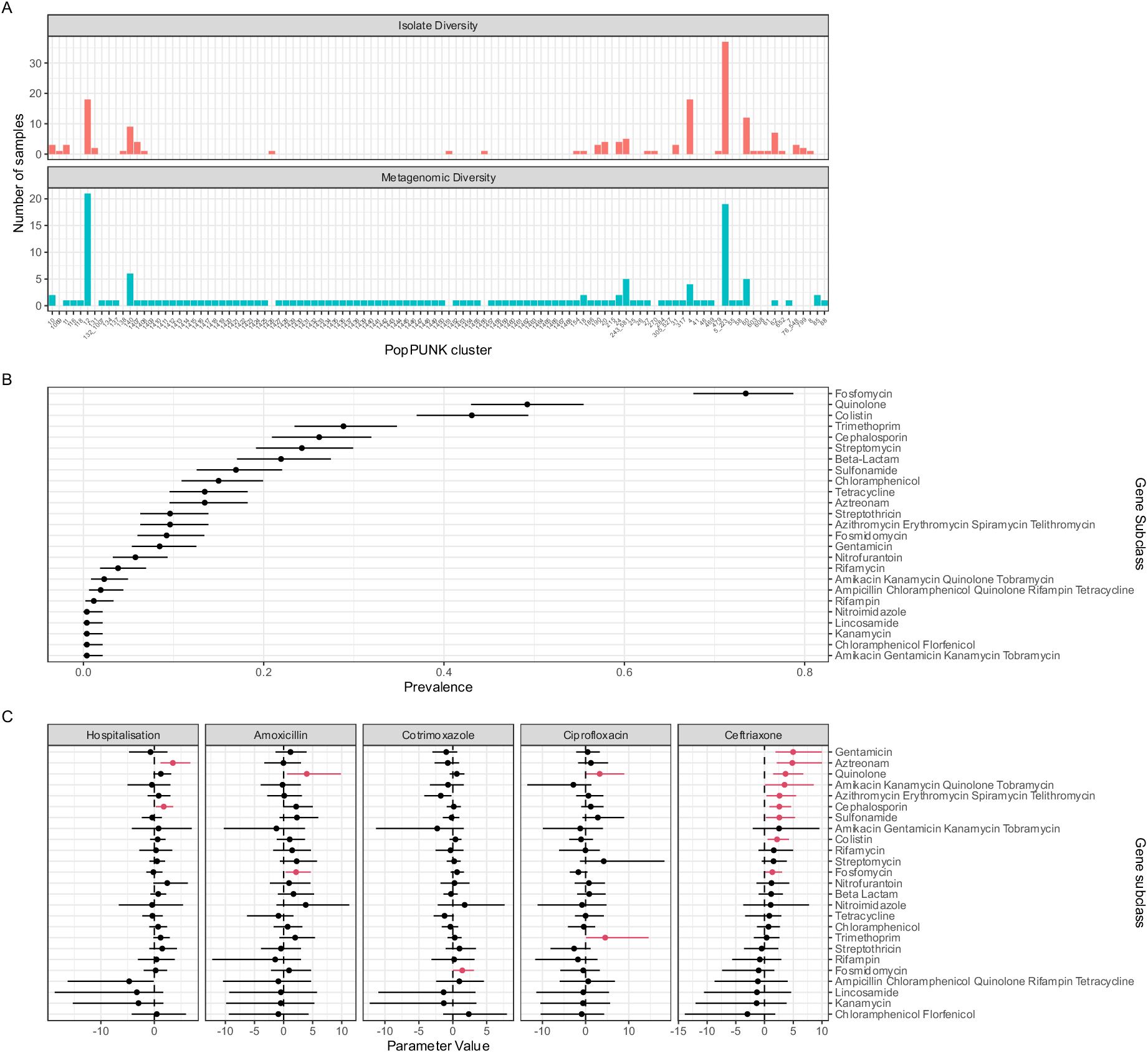
Quantifying resistance in *E. coli*. (A) Comparing the diversity of the metagenome-assembled *E. coli* genomes to genomes from ESBL-selective culture using popPUNK; number of samples (y axis) for a given popPUNK cluster (x axis) show that the metagenome-assembled genomes are more diverse; (B) Prevalence of AMRFinderPlus gene subclass in samples with a high-quality E. coli metagenome-assembled genome (n = 259) (C) Parameter values and 95% credible intervals from fitted models quantifying the effect of exposures (panels) on the presence of AMR gene subcategory in E. coli metagenome-assembled genomes. Coefficients are on the log scale and can be interpreted as exposure resulting in a log odds ratio for the presence of a given gene. A value of 0 (dotted line) is no change.

Fitting this *E. coli* specific AMRFinderPlus subclass presence/absence data using the models described above (Figure 4, Supplementary Figure 9-10) revealed that the association of ceftriaxone exposure with aminoglycoside and macrolide resistance was at least partly mediated via resistance in *E. coli*: presence of aminoglycoside (*aac(3’)-2* and *aac(6’)-1*) and macrolide (*mphA*) genes in *E. coli* was associated (95% CrI or odds ratio > 1) with ceftriaxone exposure (Figure 4, Supplementary Figure 10). Ceftriaxone exposure with AMR determinants of multiple other subclasses including cephalosporin (largely *bla*_CTX-M-15_ and *bla*_CMY2_ genes), but also of the quinolone (largely QRDR point mutations) and sulphonamide (largely *sul* alleles) subclasses, and point mutations associated with fosfomycin, colistin and aztreonam resistance (Figure 4, Supplementary Figure 10). Quinolone exposure was convincingly associated with presence of quinolone and trimethoprim resistance determinants in *E. coli*; amoxicillin exposure with quinolone and fosfomycin resistance mutations in *E. coli*, whereas cotrimoxazole exposure was not associated with clinically relevant antimicrobial resistance determinants in *E. coli*. Hospitalisation, independent of exposure to antimicrobials of these four classes, was associated with presence of resistance determinants of the cephalosporin and aztreonam subclass in these *E. coli* genomes.

## Discussion

We present here a description of the bystander effect of antimicrobial treatment for sepsis in Blantyre, Malawi on microbiome and resistome composition. Using a Bayesian modelling approach we quantify this effect, finding that different antimicrobials act to promote different bacterial taxa and resistance genes. Ceftriaxone, the first-line treatment for sepsis in Malawi, has a broad effect on microbiota and resistome composition. It is associated with an increase in abundance of Enterobacterales, including Escherichia, a key pathogenic genus. It is also associated with an increased prevalence of cephalosporin, macrolide and gentamicin resistance genes in *E. coli*, and non-Bacteroides cephalosporin, macrolide, gentamicin and rifamycin resistance gene prevalence generally in the resistome. Ciprofloxacin is similarly associated with an increase in Enterobacterales, with an increase in quinolone resistance prevalence in *E. coli* and macrolide and aminoglycoside resistance gene prevalence in the resistome. We did not identify a clear association of ciprofloxacin exposure with cephalosporin resistance, but the relative rarity of ciprofloxacin exposure meant that confidence intervals were wide. Amoxicillin and cotrimoxazole had a less pronounced effect on microbiome and resistome composition though again credible intervals were wide for these agents. Importantly, non-Bacteroides cephalosporin and beta-lactam resistance genes were associated with cotrimoxazole exposure, an important finding when cotrimoxazole is used at huge scale in community settings across the country in the context of cotrimoxazole preventative therapy in HIV. These gross antibiotic-specific perturbations in microbiome and resistome return to baseline over a timescale of around a month, though there is evidence of persistent changes in composition (beta diversity compared to baseline) out to six months. We can draw several conclusions from our findings.

First, and most importantly, quantification of the bystander effect on resistome provides evidence to inform antimicrobial stewardship protocols in Malawi and elsewhere. One of the aims of stewardship is to minimise antimicrobial exposure (both in terms of duration and spectrum), but without a quantitative measure of the off-target effect of individual antimicrobial agents, such strategies cannot be fully evidence based. Ceftriaxone, the first-line treatment for sepsis in Malawi, demonstrates a profound effect on microbiota and resistome composition but these deleterious effects must be balanced against its activity on the locally prevalent pathogens. By quantifying this effect - using simulation from the posterior of our fitted models to explore counterfactuals – our approach can inform this decision.

Second, our findings highlight important similarities but also differences between our cohort and high-resource settings. A reduction in microbiome diversity in high-resource settings following antimicrobial exposure is well described, both in healthy volunteers^9,15,16^, and hospitalised inpatients^17,18^ and most studies report an approximate return to baseline alpha diversity over months^9,15^, though differences in species or resistome composition or pre-to-post treatment beta diversity may persist for six months of more, as we demonstrate. In our analysis, we link antimicrobial exposure to presence of Proteobacteria, particularly Enterobacterales. Elsewhere, blooms of pathobionts including *E. coli* and *Klebsiella* spp. have been described in healthy volunteers following meropenem/vancomycin/gentamicin administration in the US^15^, but in Oxford, UK^18^ ceftriaxone and ciprofloxacin exposure were associated with *reduced* Enterobacterales abundance in hospital inpatients. A potential explanation could be a difference in resistance mechanisms present in commensal *E. coli* circulating in the community between UK and Malawian settings: we have previously demonstrated very high prevalence of human carriage and environmental contamination with ESBL producing *E. coli* and Klebsiella pneumoniae in our setting in Malawi^14,19^.

Directly comparative data linking antimicrobial agent to resistance gene presence from elsewhere are scanty. Previous studies in high-resource settings have linked some individual agents (e.g. meropenem, vancomycin, gentamicin, levofloxacin, azithromycin, cefpodoxime) to changes in diversity of composition and resistome, often in healthy volunteers and mostly using aggregate measures of resistome diversity^9,15,17,20^. Heterogeneity in analysis makes it difficult to compare between studies or extrapolate from the findings to the impact on a given antimicrobial stewardship strategy in different geographical settings. Nevertheless, where sought, a bystander effect on resistome is generally demonstrated: Cefpodoxime (a third generation cephalosporin) was found to increase the Bacteroides-associated beta-lactamase gene *cfxA* and tetracycline resistance genes *tetO* and *tet40* in 20 healthy volunteers in the United States^15^. A patient cohort in Oxford, UK^18^ found an increase in abundance of aminoglycoside and tetracycline resistance genes following beta lactam exposure (as we demonstrate). In Germany ciprofloxacin was associated with an increase in macrolide and cephalosporin resistance gene abundance but a *decrease* in aminoglycoside resistance gene abundances (the opposite of our findings), compared to cotrimoxazole^15^. This highlights the complexity of the antimicrobial effect on microbiome which is likely modified by diet, pretreatment bacterial structure, and host living environment including water sanitation and hygiene.^21,22^

Third, our findings highlight a key role for *E. coli* as a carrier of ARG in our setting; antimicrobial exposure (particularly ceftriaxone and ciprofloxacin) acts to increase abundance of Enterobacterales, (which seems to be largely due to Escherichia abundance) and drug resistance in *E. coli.* In high income settings, *E. coli* has repeatedly been identified as a significant contributor to ARG carriage in the microbiome of infants and young adults^6,20^. Presence of drug-resistant *E. coli* in stool increases risk of subsequent infection^23^ and microbiome-modulating strategies such as faecal microbiota transplant have been demonstrated in case reports to promote colonisation resistance to resistant Enterobacterales^24^. Given our findings, strategies to reduce colonisation with drug-resistant Enterobacterales could have a significant role to play in moderating the resistome in people treated with antibiotics in Malawi, with a subsequent effect on invasive infection. This would certainly include improved water, sanitation and hygiene in the community and infection prevention and control strategies in hospital, but also potentially trials of probiotic/prebiotic and antimicrobial stewardship with bystander AMR as an outcome.

The strengths of our study are our longitudinal sampling, and modelling approach, which together allow us to account for between-person microbiome variation. Our Bayesian modelling strategy allows us to fit complex models in a computationally straightforward manner (allowing the timescale of decay of antimicrobial effect to vary, for example) and to move beyond null hypothesis significance testing and simulate from the posterior of our models to explore the effect of different antimicrobial strategies. The flexible modelling approach can be easily modified to reflect different sampling strategies. The major limitations of the study are that, despite the modelling, residual confounding is very likely to remain which warrants caution in causal interpretations: for example, cotrimoxazole is largely used as a preventative therapy which is associated with HIV. Given the collinearity of HIV/CPT, it is not possible to include both variables in the models. Some exposures – ciprofloxacin and particularly amoxicillin, were rare compared to ceftriaxone, resulting in wide confidence intervals of parameter estimates, so absence of evidence of an effect may not represent evidence of absence. These effects are important to quantify in the future given the importance of these agents in the community. We present simulations from the posterior but have not carried out any formal out-of-sample validation, hence the models are likely overfit to data and should not be interpreted as predictions but rather aids to understanding the effects of different covariates in the models. True predictive modelling would require a larger data set. Though we present an analysis linking antimicrobial exposure to microbiome changes, the link between these changes and clinically important outcomes - transmission, and development of invasive infection - are not well understood and should be a priority for future research. In keeping with other short-read shotgun-metagenomic studies, assembling mobile genetic elements and linking them to the bacterial genomes with which they were associated is not possible; it is likely therefore that our *E. coli* binning approach may underestimate *E. coli* associated AMR if MGEs are not correctly assigned to a bin.

In conclusion, we use metagenomic sequencing and Bayesian modelling to quantify the bystander effect of antimicrobials on microbiome composition and resistome in Malawi. We demonstrate strong promotion of Proteobacteria, particularly Enterobacterales, associated with ceftriaxone and ciprofloxacin exposure, and off-target increase in prevalence of aminoglycoside and macrolide resistance genes. These changes are mediated at least in part by presence of resistance genes in *E. coli*. Amoxicillin and, particularly, cotrimoxazole, are associated with less microbiome and resistome perturbation, though cotrimoxazole shows an association with cephalosporin resistance which, given the widespread national use of cotrimoxazole preventative therapy, may be a significant driver of ESBL gene colonisation. These findings can begin to inform antimicrobial stewardship protocols in Malawi.

## Supporting information

Sample ENA accession numbers

## Online Methods

### Study design and setting

The design of the observational clinical cohort study on which this analysis is based is described in detail elsewhere^13^. The study was approved by the research ethics committees of the Liverpool School of Tropical Medicine (16-062) and Malawi College of Medicine (P.11/16/2063).

Samples were collected between February 19th 2017 and 1st May 2019. In arm one, adults > 16 years old with sepsis - defined as fever (or history of fever within 72 hours) and evidence of organ dysfunction (oxygen saturation <90%, systolic blood pressure

<90mmHg, respiratory rate > 30 breaths/minute or Glasgow Coma Score <15) were enrolled. Two comparator cohorts were also enrolled: arm 2 was composed of age- and sex-matched adults being admitted to hospital via the Emergency Department but with no current plan for antibiotic therapy from the attending clinical team, and arm 3 was a group of age-, sex- and location-matched community controls. Participants were excluded from the comparator groups if they had received antibiotics within the past four weeks. Hospitalised participants were followed up daily by a member of the study team to record exposure to antibiotics. Treatment decisions were taken by clinicians who were independent from the study team. On discharge, patients were followed up at approximately days 7, 28, 90 and 180, corresponding to visits 1, 2, 3 and 4. Day 7 and 90 (1 and 3) visits were omitted for the community group. At baseline (visit 0) and at each study visit, a stool sample or rectal swab was collected. Following incubation in enrichment broth, culture and identification (using analytical profile index) for the presence of ESBL-E organisms was carried out, and primary samples were stored.

### Isolate sequencing

Isolates selected for whole genome sequencing and their extraction have been described previously^25,26^, and these data are available at in the European Nucleotide Archive under project IDs PRJEB26677, PRJEB28522, PRJEB36486 and PRJNA869071 with linked metadata available as the R *blantyreESBL* package^27^. Briefly, DNA was extracted from overnight nutrient broth cultures using the Qiagen DNA Mini Kit (Qiagen, Germany) according to the manufacturer’s instructions. Sequencing was performed at the Wellcome Sanger Institute using the Illumina HiSeq X10 system (Illumina Inc., USA), producing 150 bp paired-end reads.

### Metagenomic sequencing

A subset of 450 stored primary samples from 163 participants were selected for metagenomic sequencing. These were selected on pragmatic grounds to maximise longitudinal representation. Samples were not sequenced from individuals who had only completed one study visit. Due to the process of selecting samples for sequencing, age- and sex-matching of the comparator cohorts was not maintained. Genomic material was extracted using the Qiagen DNA stool mini kit (Hilden, Germany), with the addition of a bead-beating step. A sterile saline negative control sample for each extraction run was included (25 in total), which were also prepared and sequenced as below. Library preparation was performed with the NEBNExt Ultra

II FS kit (New England Biolabs, Ipswich, Massachusetts, Unites States) on the Mosquito Ultra II platform (Qiagen), using a 1/10 reduced volume protocol. Metagenomic sequencing was performed at the University of Liverpool Centre for Genomic Research (Liverpool, U.K), using the Illumina NovaSeq platform with an S4 flow cell (San Diego, California, United States). Sequencing was multiplexed and aimed at a depth of 100 million reads per sample. Reads were deposited in the European Nucleotide Archive, under project accession PRJEB86881.

## Bioinformatic analysis

### Quality control of reads

Modules from the MetaWRAP (v1.3.2) pipeline^28^ were used to standardise metagenome analysis. All paired-end reads underwent quality-control using the MetaWRAP “read_qc” module to remove low-quality, adapter, and human sequence reads. The T2T consortium complete human genome, (GCF_009914755.1) and human mitochondrial genome (NC_012920.1) were used as references for the removal of human reads. Samples with sequencing failure (fewer than 0.5million reads were excluded from further analysis). The remaining samples had a median (IQR) 129 (97-147) million reads per sample.

### Read-based taxonomy

Reads were assigned taxonomy with Kraken2^29^ (v2.1.2) using a custom database comprising all RefSeq complete genomes and proteins for archaea, bacteria, fungi, viruses, plants, and protozoa. This database also included complete RefSeq plasmid nucleotide and protein sequences, as well as a version of the NCBI UniVec database minimised for false positives. A confidence threshold of 0.1 was applied for read assignments, and reports were generated for downstream analysis.

### Assembly of metagenomes and *E. coli* bins

Metagenomic reads were assembled using metaSPAdes^30^ (v3.1.3) under default settings. To generate the best bins possible, three binning strategies (MetaBAT2, CONCOCT and MaxBin2) were employed across all samples via MetaWRAP. The reference genome for the *E. coli* type strain (ATCC 11775, Accession: GCF_003697165.2) was then used as a reference for FastANI^31^ (v1.3.3) to identify any bins that were *E. coli* to the species level (>95% ANI). CheckM2^32^ (v1.0.2) was then used to select bins that meet MIMAG standards (>50% completion, <10% contamination). For each sample, the most complete bin was then selected for downstream analysis, yielding 261 bins, representing the same number of samples.

### Calling antimicrobial resistance from metagenomes and quantifying the bystander effect

To understand the general contribution of the microbiome to antimicrobial resistance, metagenomes were analysed to find acquired antimicrobial resistance genes using AMRFinderPlus^33^ (v3.11.20) under default settings. Once *E. coli* was determined to be a primary organism of interest, a second analysis was conducted using an *E. coli*-specific model within AMRFinderPlus, allowing detection of point mutations associated with resistance. This refined analysis was applied to the metagenome, *E. coli* genome bins, and *E. coli* isolates. This approach aimed to compare the diversity of AMR genes between cultured and uncultured samples and to quantify the extent to which specific AMR determinants could be attributed to the *E. coli* metagenomic bins. Isolate reads were assembled using SPAdesSPA^34^ (v3.1.3).

### Cultured vs uncultured diversity using PopPUNK

In order to characterise diversity in a comparable way, PopPUNK^35^ (v2.7.2) was used to assign clusters to E. coli bins and isolates, querying the “reference only” *Escherichia coli* v2 database. The resulting PopPUNK clusters were visualised in Microreact^36^ and summarised as frequency tables to identify the dominant clusters observed when comparing metagenomic versus culture-based approaches.

### Statistical Analysis

All analysis was carried out in R^37^ v4.4.2 and all plots were generated with ggplot^38^ v3.5.1. Unless otherwise stated, summary statistics are presented as medians with interquartile ranges or proportions with exact binomial confidence intervals. Shannon Diversity and Bray-Curtis Dissimilarity were used to quantify alpha and beta diversity, and principal components analysis on alpha diversity were carried out using the PhyloSeq^39^ v1.5.0 R package. Kruskall-Wallace test to compare Bray-Curtis dissimilarity to baseline across the three study arms was carried out with ggstatsplot v0.13.0.

### Linear regression: Shannon Diversity

To quantify the effect of antimicrobial exposure on Shannon diversity, taxa abundance and presence of AMR genes, we constructed Bayesian regression models using the Stan probabilistic programming language, accessed via cmdstanR v0.8.1 using cmdstan v2.35. All the models had the same correlation structure to account for repeated measurements on individuals; the simplest model with the linear regression model for Shannon diversity, where the sample *y*_*y*_ is given by

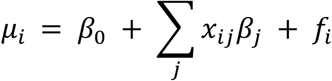

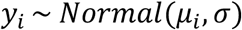

Where μ is the mean and σ the standard deviation of the normal distribution. *i* = 1,2, … *n* where *n* is the number of samples. There are *j* covariates in the model; *x_ij_* is a matrix with *n* rows: row *i* gives the covariate values for sample *y_i_*; β*_j_* are the regression coefficients. *f* encodes the within-participant correlation; it is 0 for different participants, but within-participant is defined by

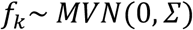

i.e. a multivariate normal distribution with a k-by-k covariance matrix for *k* within-participant samples. This matrix uses an exponentiated quadratic kernel:

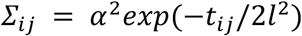

Where *t_ij_* is the difference in time between two samples. α encodes the magnitude of the within-particpant correlation and *l*, the length scale, the temporal correlation.

The effect of antimicrobials was encoded in a time-dependent manner, β(*t*).

If antimicrobials are administered between *t_a_*_’_ and *t_b_* then during exposure, the effect of antimicrobials is

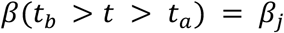

The effect after exposure is

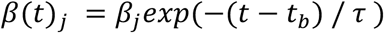

And the effect before any exposure (or for a participant with no exposure) is

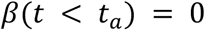

This allows the effect of antimicrobial exposure to decay exponentially on a time scale fit to the data.

Priors were student t distribution with 3 degrees of freedom and mean 0, sd 3 for β, τ, α. *l* used a inverse gamma distribution with both parameters set to 3; the mean of β_0_ was 3.

### Negative binomial regression: taxon abundance

Modelling absolute read numbers assigned to a given taxa used the same linear predictor, but assumed that the read number was negatively binomially distributed with an over dispersion parameter φ, and added an offset to account for varying read depths

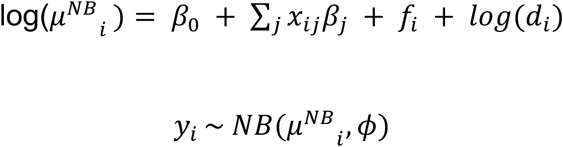

Where *d*_*y*_ is the number of reads for sample *i*, and *y_i_* here is the number of reads assigned to the taxa of choice. A different model was fit for the top 10 phyla, orders, and genera. Priors were the same as the linear model except the intercept (β_0_) was set to a mean of -3.

### Logistic regression: AMR gene presence

Modelling presence or absence of AMR gene used the same linear predictor as above, but a logistic regression model and logit link function

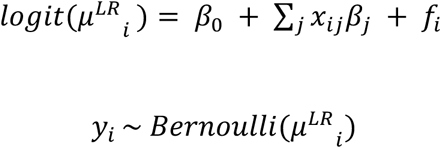

Priors were the same as the linear model except the intercept (β_0_) was set to a mean of 0.

### Fitting and simulating from the models

Time was scaled by the standard deviation. All models were run with 4 chains for 1000 iterations with a warmup of 500 iterations. Convergence was assessed by the Gelman-Rubin statistic being close to 1, and inspection of traceplots. Posteriors were summarised using median and 95% credible intervals, unless otherwise stated. For simulations, covariate values were fixed and the whole posterior (excluding warmup) used to generate predictions of the population mean outcomes of interest (i.e. ignoring within-participant correlations), which were then summarised with medians and 95% quantiles, unless otherwise stated.

## Data availability

Metadata for the participants from whom samples were collected an including antimicrobial exposures are available via the *blantyreESBL* v1.4.1 R package^27^ accession numbers of individual samples are in Supplementary Data, linked back to metadata by the lab_id variable. All data and code necessary to replicate the analyses here are available at the project github repo, https://github.com/joelewis101/deep_sequencing.

## Funding

This work was partly funded by a Wellcome Clinical PhD fellowship to J.L. (109105z/15/a) and J.L. was supported by an NIHR clinical lectureship (CL-2019-07-001). N.A.F. is funded by an NIHR Global Health Research Professorship.

E.C.-O and A.C.D are affiliated to the National Institute for Health and Care Research Health Protection Research Unit in Gastrointestinal Infections at University of Liverpool, in partnership with the UK Health Security Agency (UKHSA), in collaboration with University of Warwick. E.C.-O. and A.C.D. are based at The University of Liverpool. The views expressed are those of the author(s) and not necessarily those of the NIHR, the Department of Health and Social Care or the UK Health Security Agency.

## Notes

### Competing Interest Statement

The authors have declared no competing interest.

### Author Declarations

The study was approved by the research ethics committees of the Liverpool School of Tropical Medicine (16-062) and Malawi College of Medicine (P.11/16/2063).

