## Supplementary Tables and Figures for "Quantifying the bystander effect of antimicrobial use on the diversity and resistome of the gut microbiome in Malawian adults"

### Supplementary Figures and Tables

**Supplementary Table 1:** Antimicrobial exposures in the cohort

| Antimicrobial | Number Exposed | Days of exposure (Median [IQR]) |
|---|---|---|
| Ceftriaxone | 94/162 (58%) | 5 (2-7) |
| Cotrimoxazole | 56/162 (35%) | 152 (152-152) |
| Ciprofloxacin | 30/162 (19%) | 7 (5-7.75) |
| TB therapy | 25/162 (15%) | 139 (95-149) |
| Amoxicillin | 24/162 (15%) | 6.5 (4.75-7) |
| Fluconazole | 14/162 (9%) | 5 (2-6.5) |
| Metronidazole | 13/162 (8%) | 4 (3-10) |
| Artesunate | 8/162 (5%) | 2 (1.75-3) |
| Co-amoxiclav | 8/162 (5%) | 5 (2.5-5) |
| Lumafentrine-artemether | 6/162 (4%) | 3 (3-3) |
| Doxycycline | 5/162 (3%) | 7 (6-13) |
| Azithromycin | 4/162 (2%) | 2.5 (1.75-3.5) |
| Gentamicin | 4/162 (2%) | 5 (4.75-18) |
| Erythromycin | 3/162 (2%) | 7 (6-9) |
| Aciclovir | 2/162 (1%) | 22 (15-29) |
| Flucloxacillin | 2/162 (1%) | 2.5 (1.75-3.25) |
| Chloramphenicol | 1/162 (1%) | 1 (1-1) |
| Penicillin | 1/162 (1%) | 4 (4-4) |
| Quinine | 1/162 (1%) | 1 (1-1) |
| Streptomycin | 1/162 (1%) | 8 (8-8) |

**Supplementary Table 2:** Proportion of participants with AMRFinder-defined beta-lactam subclass identified (excludes bacteroides-specific genes defined as *cfiA, cblA, crxA, cepA* or *cfxA* beta-lactamases)

| | n/N (% [95% CI]) participants with gene identified at least once | | |
|---|---|---|---|
| Class | Arm 1 | Arm 2 | Arm 3 |
| Beta-lactam | 107/109 (98% [94-100%]) | 29/29 (100% [88-100%]) | 18/24 (75% [53-90%]) |
| Carbapenem | 8/109 (7% [3-14%]) | 2/29 (7% [1-23%]) | 0/24 (0% [0-14%]) |
| Cephalosporin | 84/109 (77% [68-85%]) | 18/29 (62% [42-79%]) | 9/24 (38% [19-59%]) |

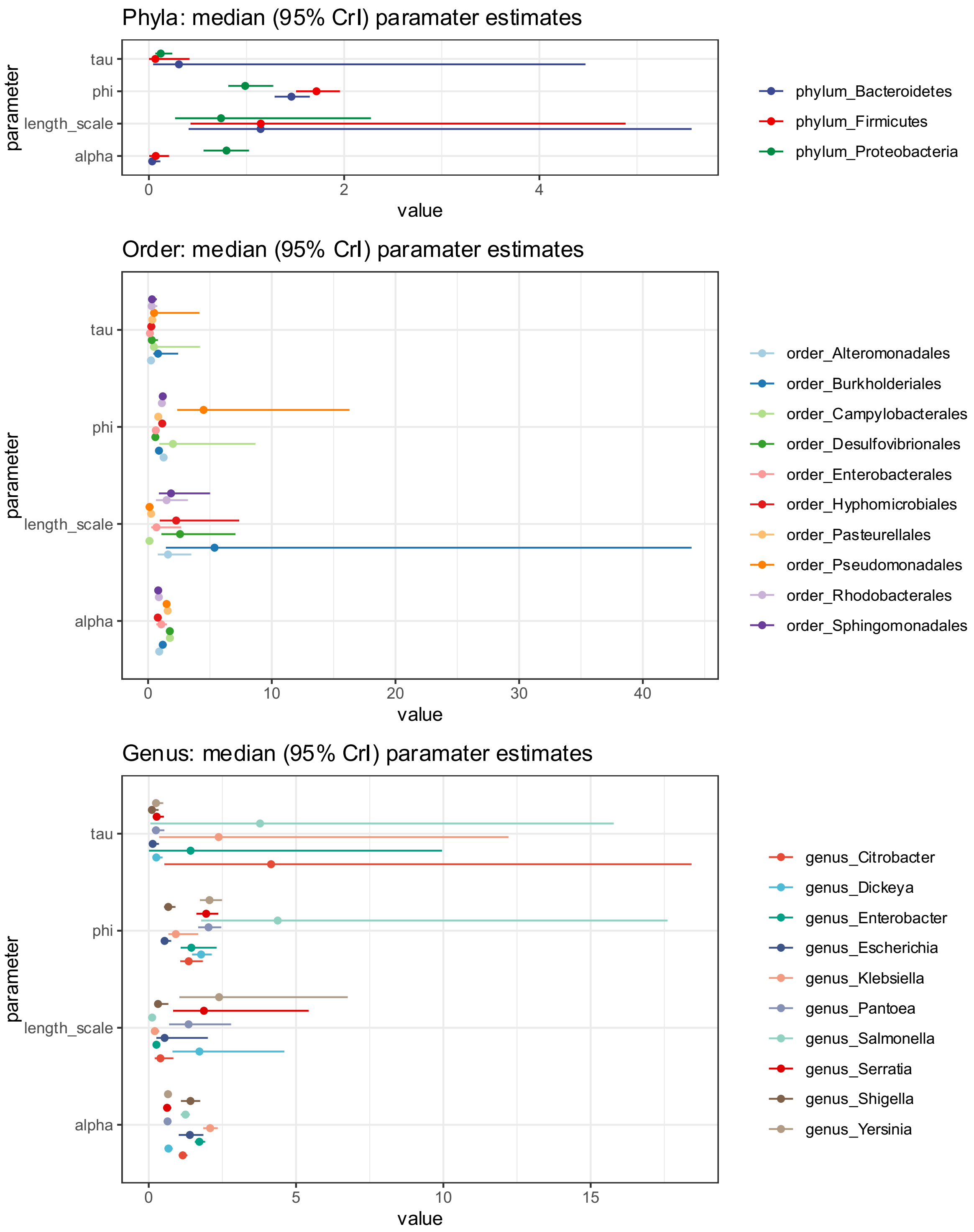


**Supplementary Figure 1:** parameter estimates (other than covariate coefficients) from fitted microbiome composition models, considering absolute read numbers assigned to a given taxa as the outcome variable. Tau is the decay constant of the exposure effect (in standardised units such that 1 unit = 55 days); phi is the overdispersion parameter from the negative binomial distribution; length scale is a measure of the within-person temporal correlation and how it decays over time; alpha is a measure of the magnitude of the within-person temporal correlation.

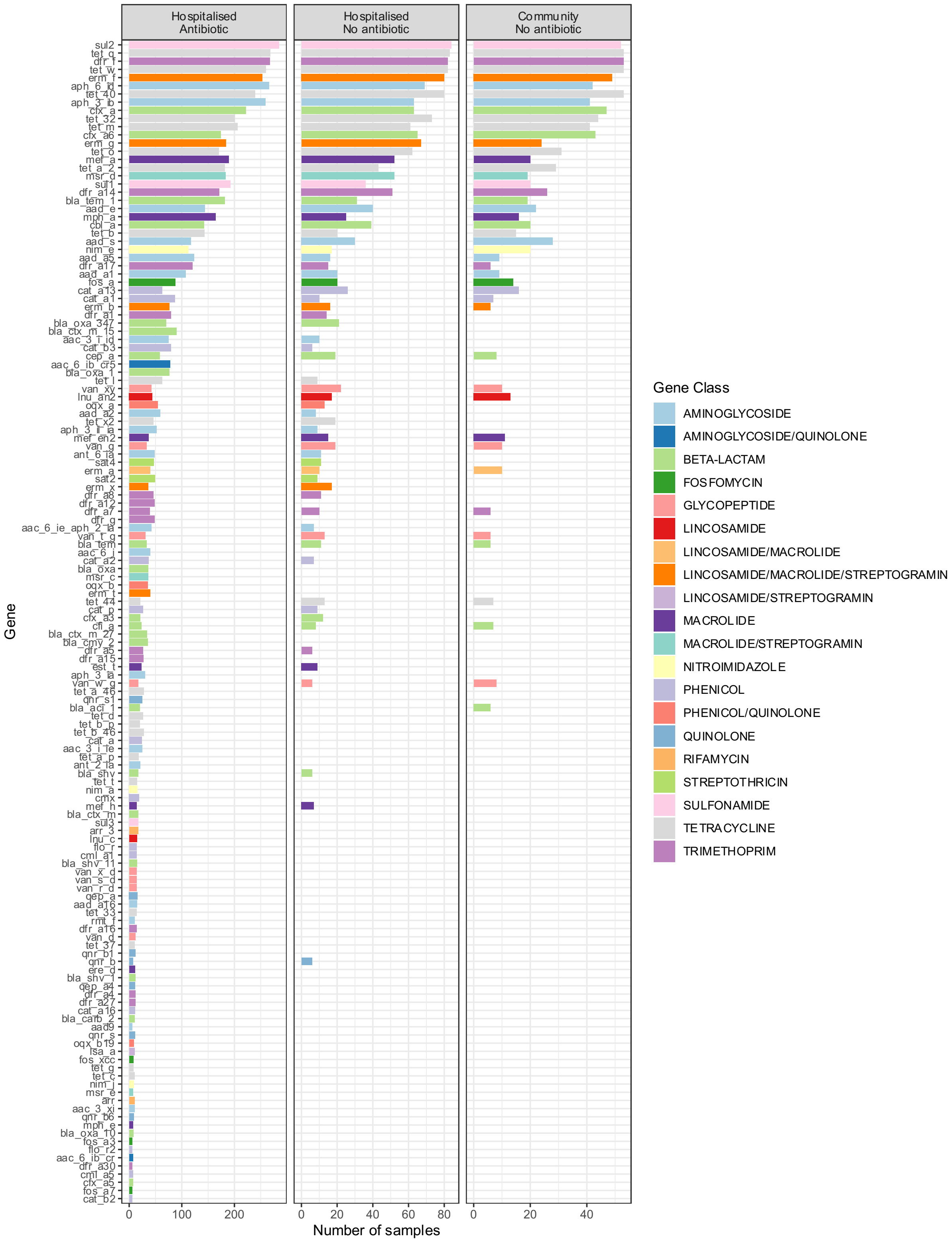


**Supplementary Figure 2:** Distribution of resistance genes identified by AMRFinder, coloured by AMRFinder-defined gene subclass, stratified by study arm.

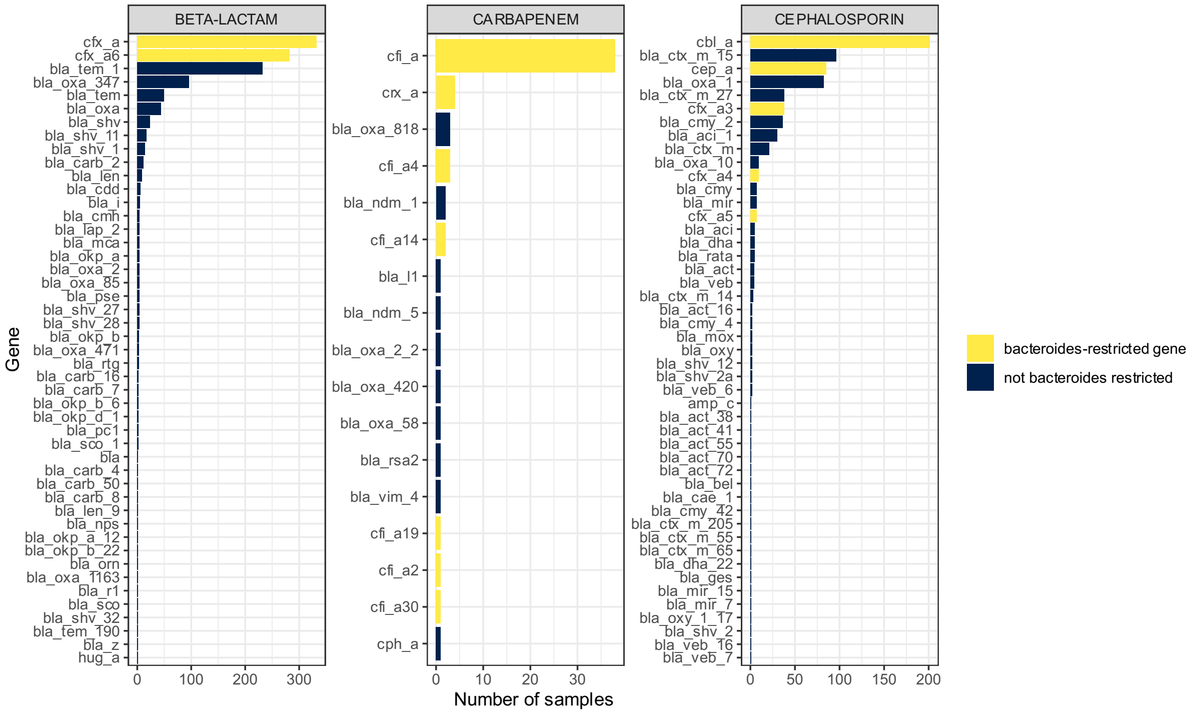


**Supplementary Figure 3:** Distribution of identified resistance genes of the AMRFinder-defined subclasses beta-lactam, cephalosporin and carbapenem, stratified by whether the gene is a bacteroidates-specific gene.

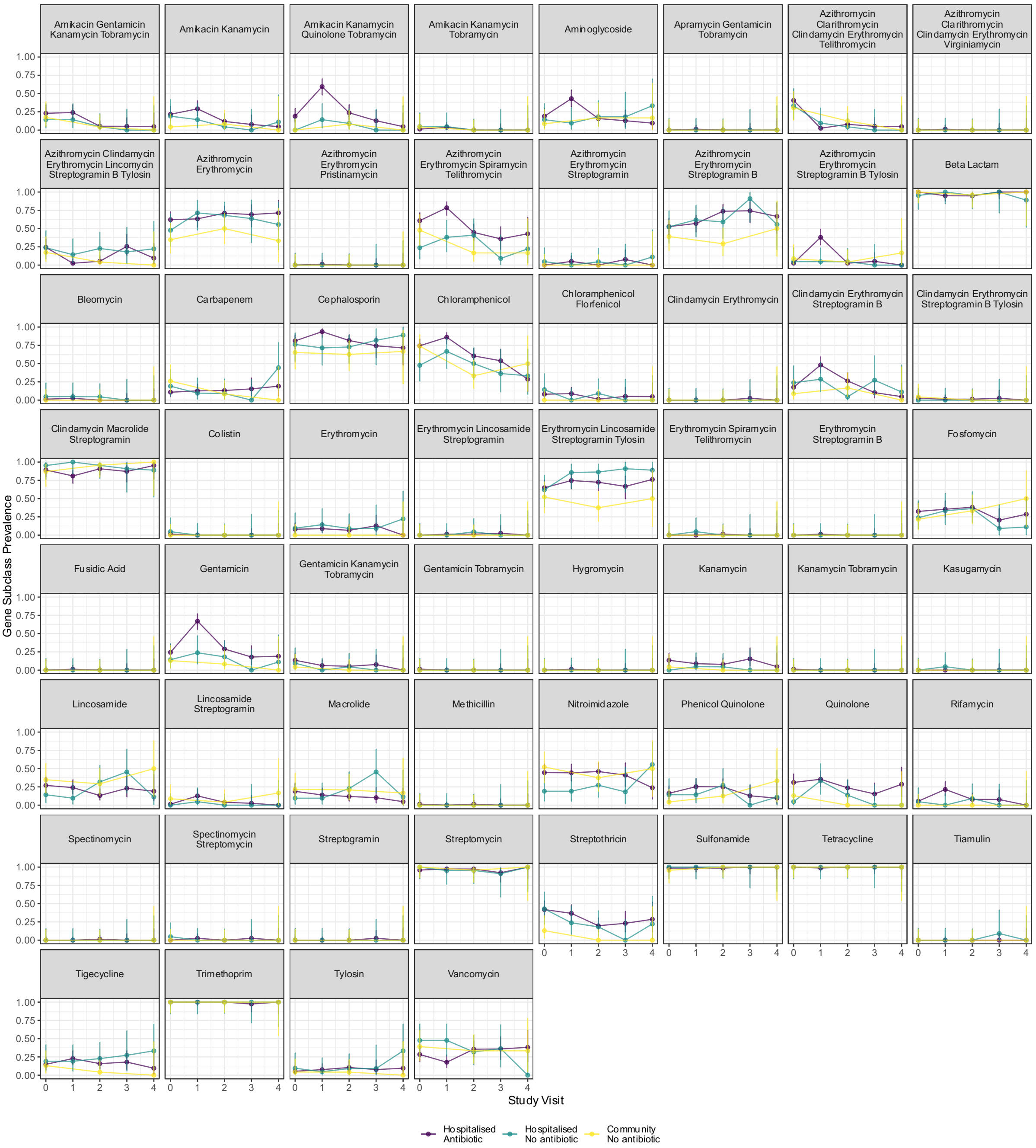


**Supplementary Figure 4:** AMRFinder-defined gene subclass prevalence stratified by study visit and arm.

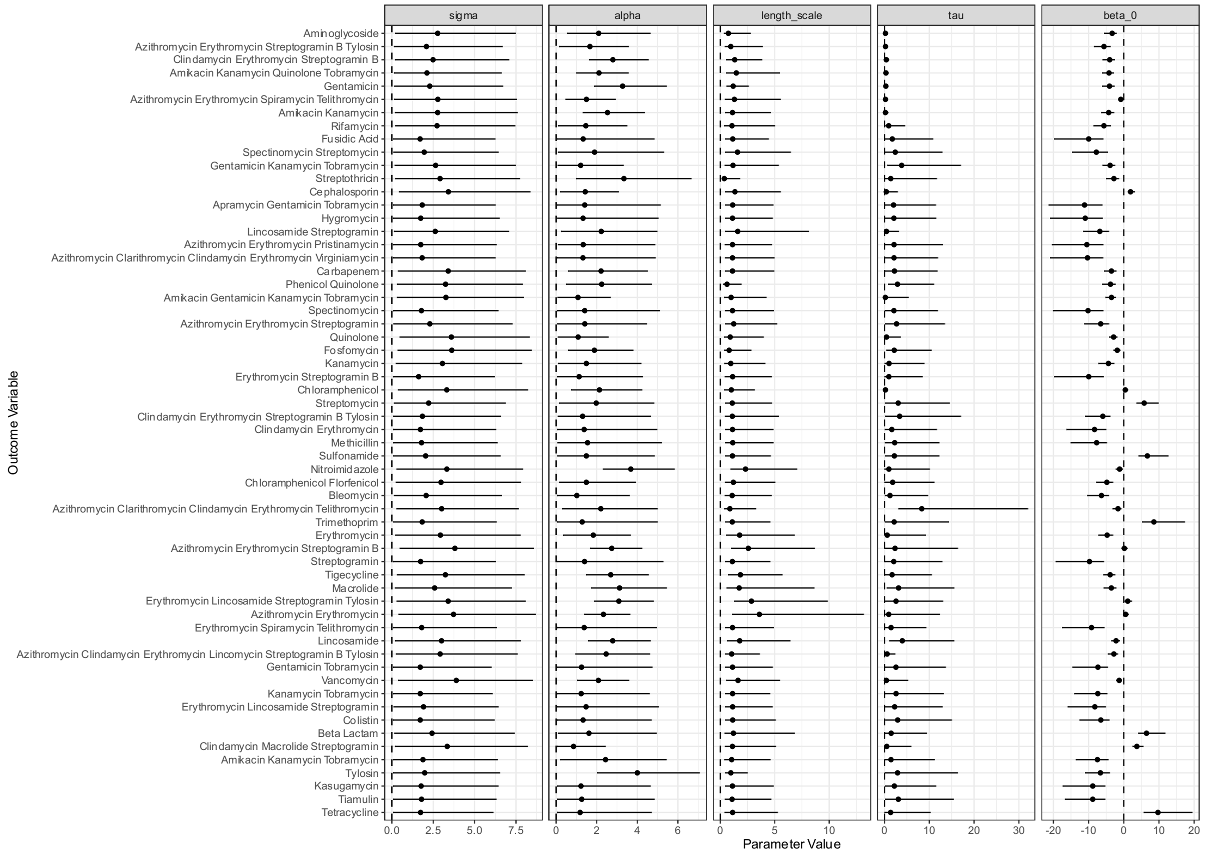


**Supplementary Figure 5:** Parameter values (x-axis) and 95% credible intervals from fitted models quantifying the effect of exposures on presence of AMR gene category (y-axis). Tau is the decay constant of the exposure effect (in standardised units such that 1 unit = 55 days); sigma is the standard deviation of the within-participant correlation at time 0; length scale is a measure of the within-person temporal correlation and how it decays over time; alpha is a measure of the magnitude of the within-person temporal correlation.

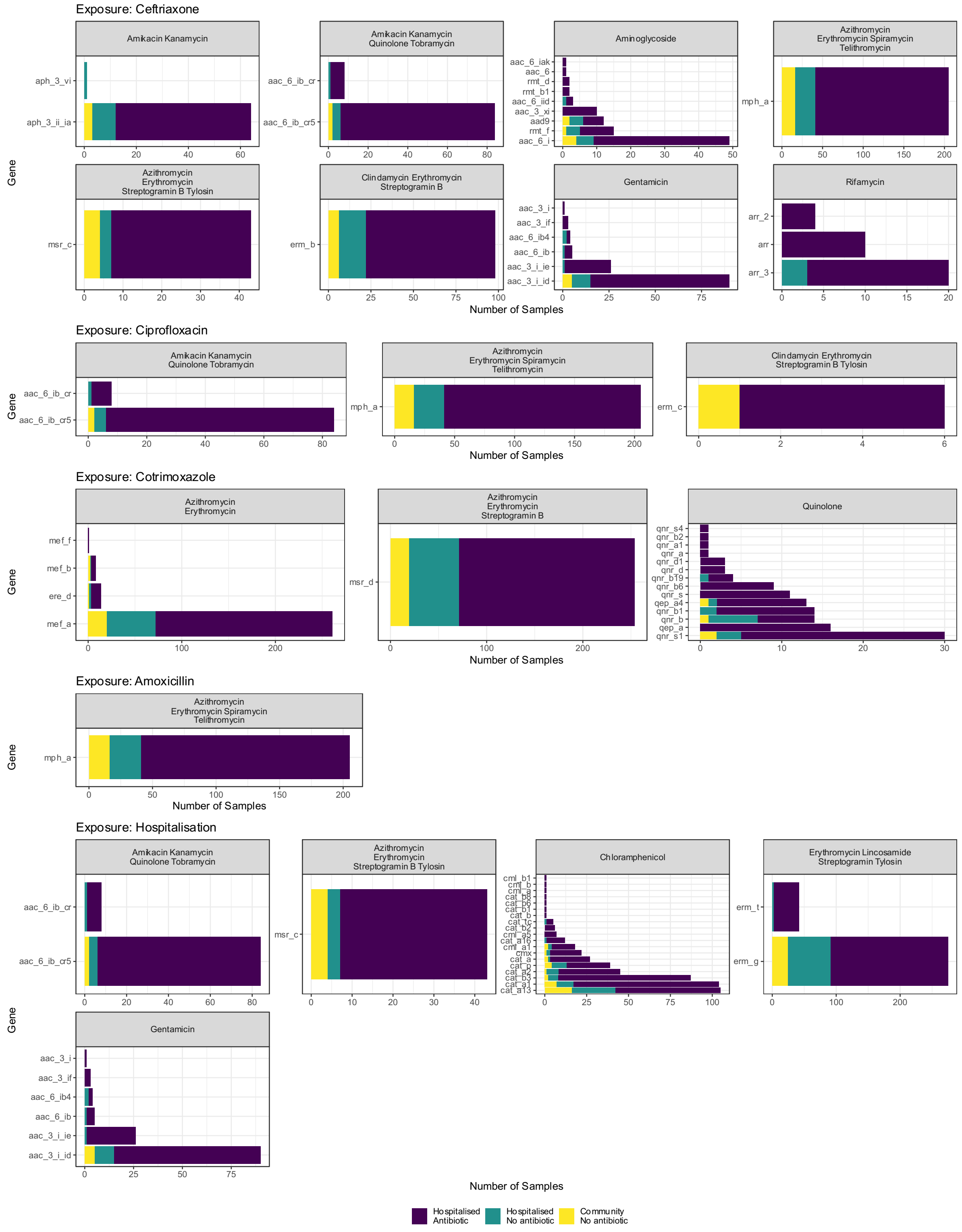


**Supplementary Figure 6:** Distribution of resistance genes in AMRFinder-defined subclasses that are increased in association with(from top) cephalosporin, ciprofloxacin, cotrimoxazole and amoxicillin exposure, and hospitalisation; increased here is defined as lower limit of 95% credible interval of odds ratio for presence of gene given exposure > 1.

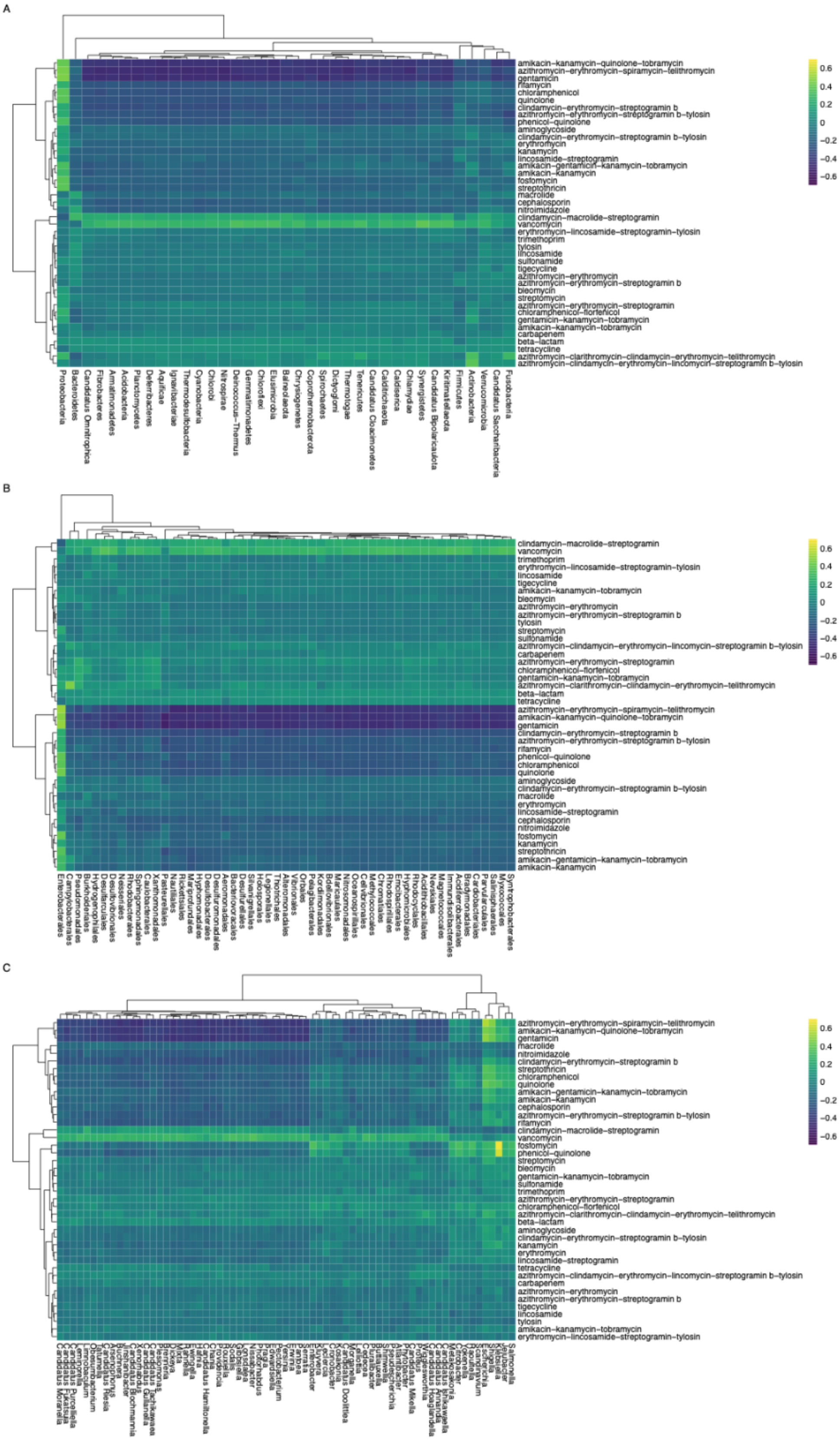


**Supplementary Figure 7:** Hierarchically clustered heatmap of spearman correlation coefficient of presence of AMR gene subclass (row) and taxon (column) - Phylum (A), Order (B), Genus (C).

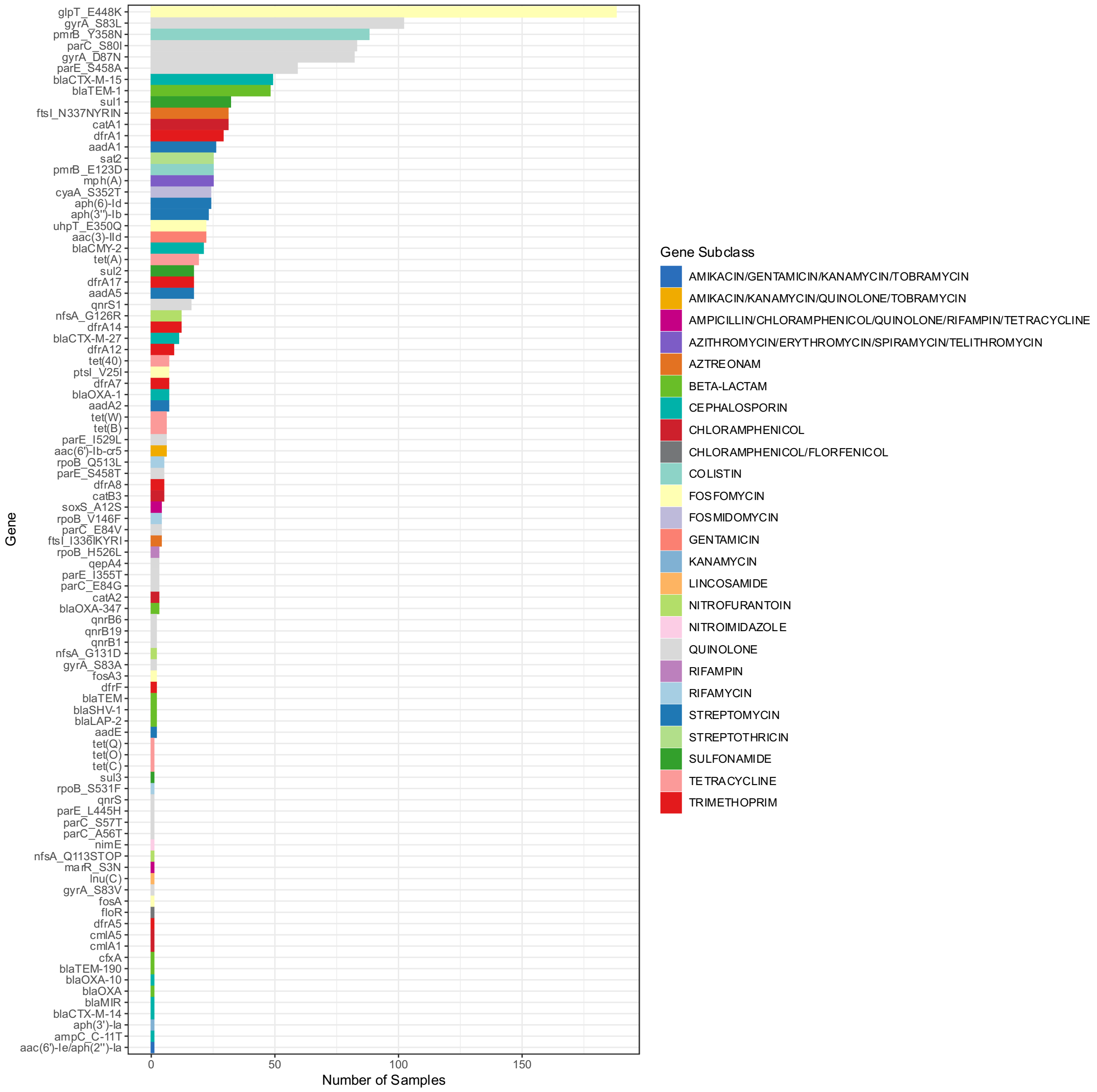


**Supplementary Figure 8:** Distribution of AMR genes/mutations identified in the E. coli metagenome-assembled genomes.

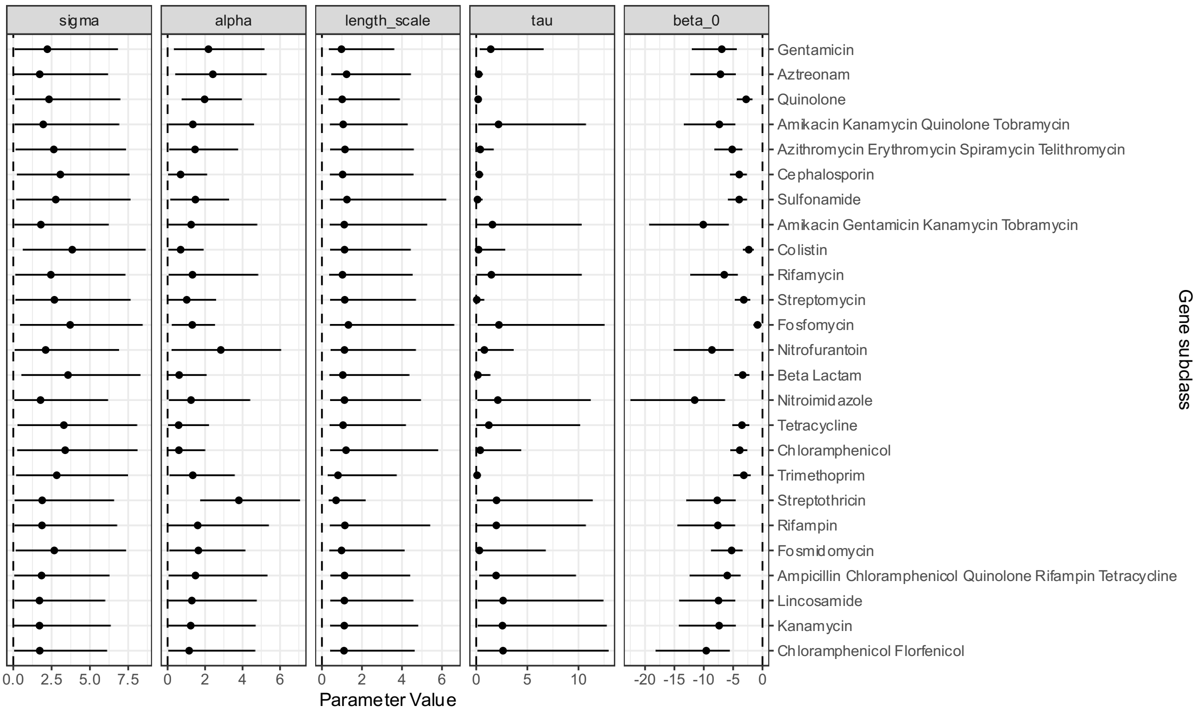


**Supplementary Figure 9:** Parameter estimates (x-axis) from models predicting presence of AMRFinder defined resistance determinants of a given subclass (y-axis) for a given parameter (panels) alpha = parameter quantifying magnitude of within-participant correlation of samples; beta_0 = intercept of linear predictor; length_scale parameter quantifying temporal association of samples within-participant; tau = decay constant of effect of covariates; sigma = standard deviation of normal distribution encoding per-participant intercept.

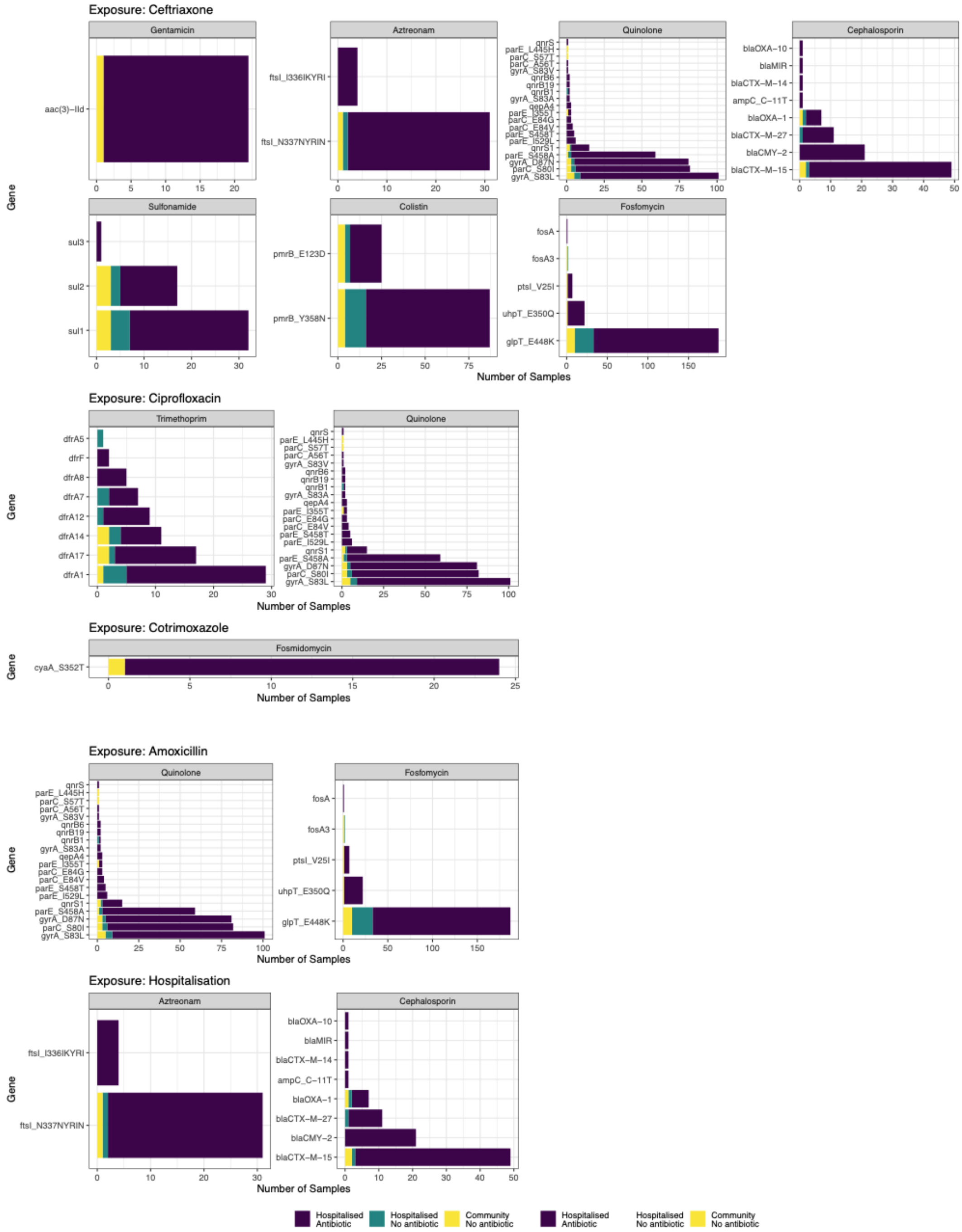


**Supplementary figure 10:** Distribution of resistance genes in AMRFinder-defined subclasses that are increased E. coli metagenome-assembled genomes in association with (from top) cephalosporin, ciprofloxacin, cotrimoxazole and amoxicillin exposure, and hospitalisation; increased here is defined as lower limit of 95% credible interval of odds ratio for presence of gene given exposure > 1.
